# A demographic adjustment to improve measurement of COVID-19 severity at the developing stage of the pandemic

**DOI:** 10.1101/2020.03.23.20040998

**Authors:** Simona Bignami-Van Assche, Daniela Ghio

## Abstract

The need for accurate statistics has never been felt so deeply as the novel COVID-19 pathogen spreads around the world and quantifying its severity is a primary clinical and public health issue. In Italy, the magnitude and increasing trend of the case-fatality risk (CFR) is fueling the already high levels of public alarm. In this paper, we highlight that the widely used crude CFR is an inaccurate measure of the disease severity since the pandemic is still unfolding. With the goal to improve its comparability over time and across countries at this stage, we then propose a demographic adjustment of the CFR that addresses the bias arising from differential case ascertainment by age. When applied to publicly released data for Italy, we show that until March 16 our adjusted CFR was similar to that of Wuhan – the most affected Chinese region, where COVID-19 has now been contained. This indicates that our adjusted CFR improves its comparability over time, making an important tool to chart the course of the COVID-19 pandemic across countries. Since March 16, the Italian COVID-19 outbreak has entered a new phase, with the northern and southern regions following different trajectories. As a result, our adjusted CFR has been increasing between March 16 and March 20. Data at the subnational level are needed to correctly assess the disease severity in the country at this stage.

## SOMMARIO

Il diffondersi dell’epidemia COVID-19 ha dimostrato l’importanza di un’informazione corretta, laddove la mancanza di una risposta tempestiva si valuta essenzialmente in termini di vite umane. In Italia, la propagazione del COVID-19 è stata accompagnata da un’informazione imprecisa circa la sua reale entità e gravità. In questo contributo, dimostriamo come il tasso di fatalità, attualmente l’indicatore più utilizzato della severità del virus, sia inaccurato a questo stadio della pandemia e proponiamo un metodo demografico per migliorarne la stima, in particolare durante la fase ascendente della curva epidemica. L’applicazione del nostro metodo ai dati nazionali italiani dimostra che, fino al 16 di Marzo, il tasso di fatalità rimane assimilabile a quello osservato nella regione di Wuhan, ma comincia a conoscere un drastico incremento negli ultimi giorni. I nostri risultati indicano che il metodo proposto migliora la comparabilità del tasso di fatalità nel tempo e tra paesi, dimostrando la sua utilità come strumento per tracciare e comprendere l’evoluzione pandemica attuale.

## Background

Italy is experiencing the worst consequences of the outbreak of COVID-19 in the world. As of March 20, the country recorded 35,731 confirmed positive cases and 3,047 deaths (*1*), surpassing the number of COVID-19-related deaths in China. The case-fatality risk^1^ (CFR), which corresponds to the proportion of COVID-19-related deaths out of confirmed positive cases, stands at 8.5% and causes concern because it has increased to this level from 2.6% on March 2. It is now substantially higher than that found in other countries, notably China where COVID-19 has been contained (*2*).

There is currently much debate about the reasons why Italy has been the country hardest hit by the pandemic, both in terms of mortality and disease severity. Although often used interchangeably, these are two distinct concepts. COVID-19-related mortality indicates the proportion of individuals in the population who die from the disease. The severity of COVID-19 measures the risk of dying among infected individuals. Overall mortality is thus determined by the severity of the disease for the proportion of infected individuals in the population, and it will be higher if disease severity is elevated for a large share of the population. This is the reason why a demographic argument frequently cited is that the old age structure of the Italian population and extensive intergenerational contact between older and younger generations have made it more vulnerable to COVID-19-related mortality (*3*). Yet this argument rests on the incorrect assumption that the disease severity is fixed and accurately measured at this stage of the pandemic, which is not the case.

Studies about novel pathogens in the past such as Ebola and SARS have shown that the during their outbreak the CFR is prone to over- or under-estimation of the real value because there is a misleading tendency to report it daily without any correction for the delay between symptoms onset and death or recovery (*4-6*). At this stage of the COVID-19 pandemic, it is not possible to assess the real value of the CFR because the final outcome (death or recovery) is not yet known for all infected cases. In addition, the majority of infections are asymptomatic or mild and do not require hospitalization thus going undetected. Therefore, corrections to the CFR are needed to improve its comparability over time and across countries.

In this paper, we argue that to achieve this goal the CFR needs to be corrected for the changing demographics of infected cases and fatalities over time. We then show how the tools of demographic analysis can be used to control for this effect, and thus improve measurement of the disease severity over time and across countries at this stage of the pandemic.

### The evolving age structure of the COVID-19 epidemic in Italy

On March 2, the Italian Health Institute (Istituto Superiore di Sanità, or ISS) received the government mandate to gather and release data about the COVID-19 epidemic. Within the scope of this mandate, ISS started publishing twice weekly a short report presenting the distribution of COVID-19 confirmed positive cases and fatalities at the national level aggregated by ten-years age groups (*7*). Our analysis uses data from the last three ISS reports, published on March 12, March 16 and March 20. Because testing procedures adopted during this period screened for COVID-19 only based on symptoms^2^, the number of infected cases largely corresponds to the number of symptomatic individuals.

Overall, the number of positive cases has increased by a factor of 2.5 between March 12 to March 20 (from 13882 to 35731). The largest increase occurred between March 12 and March 16 (from 13882 to 25058 cases), whereas the increase between March 16 and March 20 was 43%. The age pyramids of positive cases and fatalities in Figure 1 illustrate how the age structure of the epidemic has changed over this short period of time. As of March 12, the largest proportion of positive cases (40%) and deaths (90%) was found among those aged 70 years and older. This was still the case as of March 16, but the largest increase in confirmed positive cases was recorded for the age group 30-39 years (+105%), compared to 80% for 60-89 years old and 93% for individuals aged 90 years and older. Between March 12 and 20, the largest increase has also been recorded for adults aged 30-49 years, followed by the youngest (0-9) and the eldest (90+) age groups.

**Figure 1:**
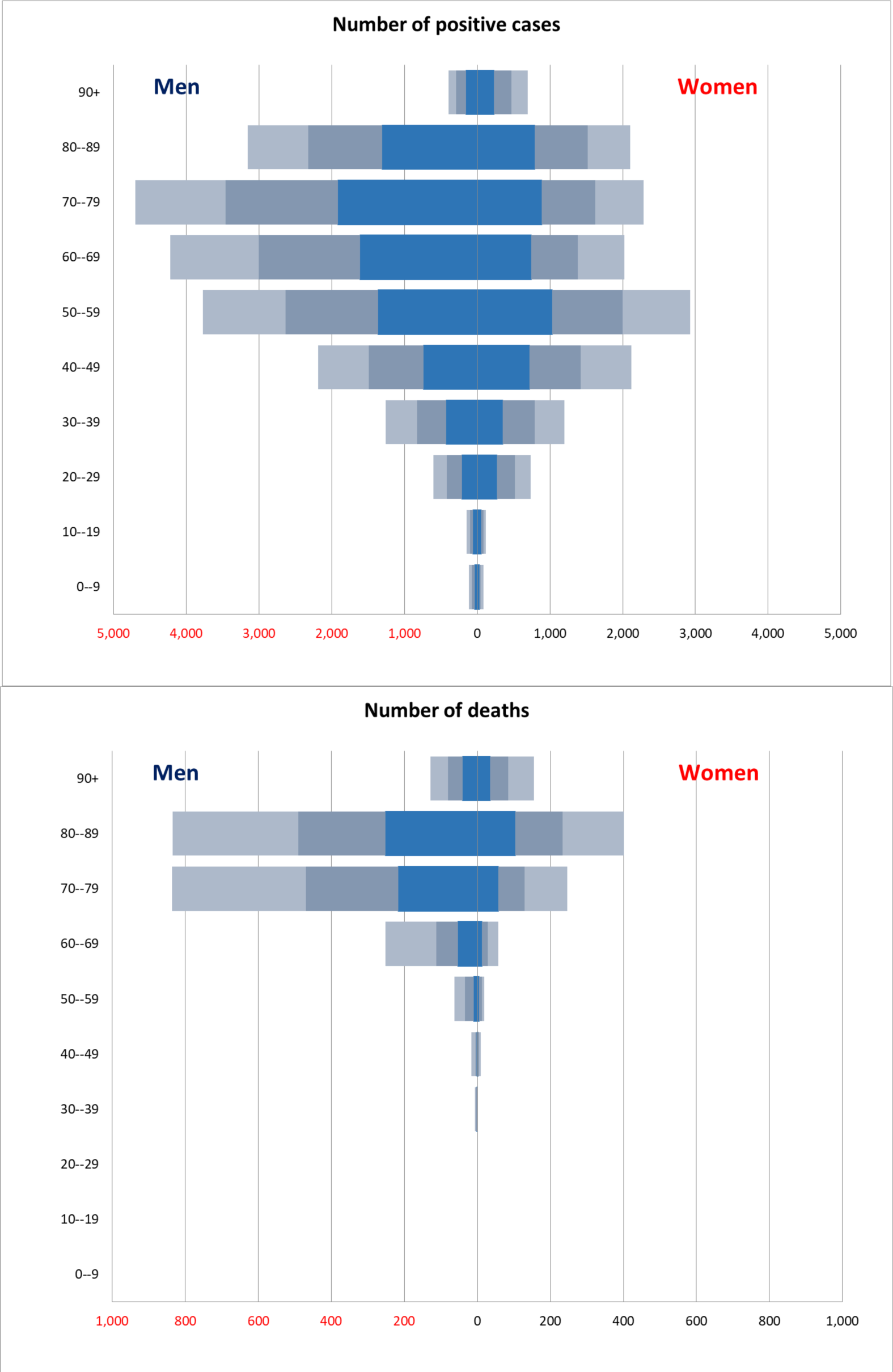
Number of COVID-19 confirmed positive cases and related deaths, by broad age groups: Italy, March 12-20 *Sources*: ISS. *Legend:* dark blue bars correspond to Italian cases and deaths recorded as of March 12; light grey bars correspond to Italian cases and deaths recorded as of March 16; light blue bars correspond to Italian cases and deaths recorded as of March 20.

The pyramids in Figure 1 also show gender differences in COVID-19 symptomatic infections. Men are more affected than women, especially in the older age groups (the upper part of the pyramid). Comparing the reported number of fatalities, the largest increase has been recorded for the age group 40-59 (267% from March 12 to March 16, 196% from March 16 to March 20). The male disadvantage observed for positive cases is confirmed for fatalities. The number of deaths among men aged 60-79 years as of March 20 is twice as many as those of women, even if the percentage increase since March 16 is the same (+170%).

The changing age distribution of COVID-19-related illness and death in Italy is the first reason why the overall CFR is an inaccurate measure to compare the disease severity over time and between populations at this stage of the pandemic. The second reason is that the number of positive cases depends on screening policies, whose procedures have also changed over time and can differ between Italian regions, and between Italy and other countries. The combined effect of these two circumstances is that the number of positive individuals *in each age group* whom are exposed to the risk of COVID-19-related death differs at each point in time and across countries. This structural effect is similar to that observed when calculating measures of fertility and mortality and it can be addressed using demographic methods.

### The adjusted case-fatality risk

Demographically, the overall CFR is a crude rate. The age structure of positive cases can be taken into account by calculating age-specific CFRs for each age group between age *a* and age *a*+*n* (CFR_*a*,,*a+n*_) and then aggregating them into an adjusted CFR (ACFR) that equals: 

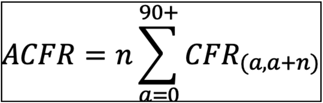

with *n* being the width of the age intervals. The ACFR can be interpreted as the average number of deaths per confirmed case of COVID-19.

We apply this adjustment to the age-specific CFRs for COVID-19 derived from the ISS data. For comparison purposes, we also apply it to the data for infected cases and deaths in the Wuhan region of China as of February 22 (*8*). As of March 12, we obtain an adjusted CFR of 4.7% in Italy (compared to a crude CFR of 5.9%), 5.7% for men (compared to a crude CFR of 7.3%) and 3.6% for women (compared to a crude CFR of 4.2%). For Wuhan, we obtain an adjusted CFR of 3.8%. The Italian ACFR was thus not much higher than the corresponding one for China. As of March 20, however, the ACFR increases substantially (to 7%), especially for men (8.2%).

**Table 1.**
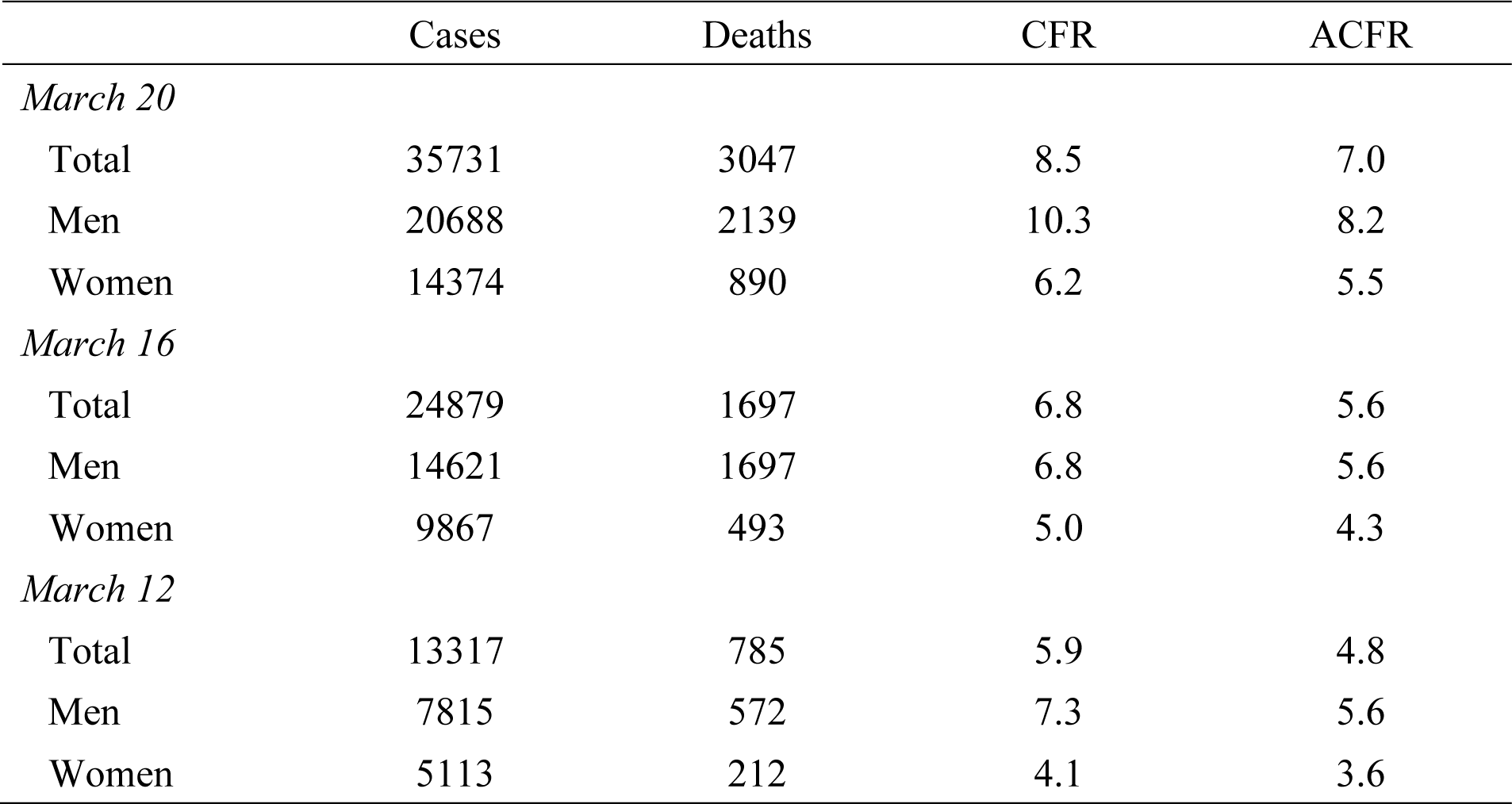
Number of COVID-19 confirmed positive cases and deaths as of March 12-20, crude case-fatality rate (CFR) and adjusted CFR (ACFR), by sex: Italy

## Discussion and conclusion

The severity profile of a novel pathogen is one of the most critical clinical and public health issues as it begins to spread, when assessing disease course and outcome is crucial for planning health interventions (*4, 5*). For this reason, the CFR is the most important epidemiological indicator to monitor during the current outbreak of COVID-19. Yet estimates of the CFR during an ongoing pandemic are biased by the calculation method. In the case of Ebola and SARS, studies have shown that crude estimates can falsely suggest that the CFR is exponentially increasing and make it difficult to understand the reasons behind differences across countries (*4-6*). In Italy, these circumstances are fueling the already high levels of public alarm and are making virologists question whether the COVID-19 infectious agent is becoming more lethal (*9*), once again as it had been the case for SARS (*10*).

The two main biases that affect the CFR during the course of an emerging epidemic are the delay between symptoms onset and death or recovery (*5*) as well as case ascertainment (*6*). The latter is a specific and important problem for the COVID-19 pandemic because, in contrast to Ebola and SARS, the majority of COVID-19 infections are asymptomatic or mild and avoid detection (*8*). In this paper, we argue that this is the main bias to be addressed to improve the comparability of the CFR over time and across countries at this stage of the pandemic. We then propose a demographic adjustment that discounts the evolving age distribution of infected cases in the calculation of the CFR. Our results show that the adjusted CFR in Italy as of March 16 was significantly lower than the crude CFR, and in a range similar to that for the Chinese region of Wuhan. This indicates that the adjusted CFR we propose could be used as a standard indicator of COVID-19 severity for comparisons across countries and over time at this evolving stage of the pandemic.

Since March 16, the Italian COVID-19 outbreak has entered a new phase, with the northern and southern regions following different trajectories. The initial outbreak that started on February 21 seems to slow down in the most severely affected northern region, but the outbreak in the central and southern region is picking up speed. As a result, our adjusted CFR has been increasing between March 16 and March 20. Additional sub-national data for cases and deaths by age are needed to better explore how local severity is contributing to the national CFR. As more data become available, it will be possible to test whether our results remain valid.

## Data Availability

Data are publicly available on the website of Italian health Institute.

https://www.epicentro.iss.it/coronavirus/sars-cov-2-sorveglianza-dati

## Acknowledgements

We would like to thank Patrick Gerland, Dessislava Choumelova, Dario Tarchi, Nikolaos Stilianakis and Ari Van Assche for their comments on earlier versions of the manuscript.

## Footnotes

1 The CFR is most-often, albeit incorrectly, referred to as the case-fatality rate or ratio. The CFR is technically a measure of risk because it corresponds to a proportion of incidence for the disease considered (the proportion of cases who die during the reference period).

2 The Italian Health System is regionalised, so that screening procedures for COVID-19 are chosen by the competent Regional Authorities. During the period we consider in the analysis all affected regions in the North of Italy, with the exception of Veneto, followed the recommendations of the World Health Organization and tested only individuals symptomatic with fever, cough, and/or difficulty breathing.

## References

1. Italian Ministry of Health. Covid-19. Situazione in Italia. www.salute.gov.it/portale/nuovocoronavirus/dettaglioContenutiNuovoCoronavirus.jsp?lingua=italiano&id=5351&area=nuovoCoronavirus&menu=vuotoDate accessed: March 21, 2020

2. Lazzerini M, Putoto G. COVID-19 in Italy: momentous decisions and many uncertainties. Lancet Glob Health. https://doi.org/10.1016/ S2214-109X(20)30110-8. Date: March 18, 2020. Date accessed: March 19, 2020.

3. Dowd JB, Rotondi V, Andriano L et al. Demographic science aids in understanding the spread and fatality rates of COVID-19. https://osf.io/se6wy/?view_only=c2f00dfe3677493faa421fc2ea38e295 Date: March 15, 2020 Date accessed: March 19, 2020.

4. Ghani AC, Donnelly CA, Cox DR et al. Methods for Estimating the Case Fatality Ratio for a Novel, Emerging Infectious Disease. American Journal of Epidemiology 2005; 52(5): 479–486.

5. Lipsitch M, Donnelly CA, Fraser C et al. Potential Biases in Estimating Absolute and Relative Case-Fatality Risks during Outbreaks. PLoS Negl Trop Dis 2015; 9(7).

6. Wu JT, Leung K, Bushman M et al. Estimating clinical severity of COVID-19 from the transmission dynamics in Wuhan, China. Nat Med 2020. https://doi.org/10.1038/s41591-020-0822-7 Date: March 19, 2020 Date accessed: March 21, 2020

7. Istituto Superiore di Sanità. Sorveglianza Integrata COVID-19: i principali dati nazionali. https://www.epicentro.iss.it/coronavirus/sars-cov-2-sorveglianza-dati Date: March 16, 2020 Date accessed: March 18, 2020

8. Novel Coronavirus Pneumonia Emergency Response Epidemiology Team. The epidemiological characteristics of an outbreak of 2019 novel coronavirus diseases (COVID-19) in China. Chinese Center for Disease Control and Prevention Weekly 2020; 41:145–51.

9. Il Giornale. In Italia il virus é più letale: ora la mortalità é salita al 9%. https://www.ilgiornale.it/news/cronache/coronavirus-italia-tasso-letalit-salito-9-1844455.html Date: March 21, 2020 Date accessed: March 22, 2020

10. Cable News Network (CNN). SARS becoming deadlier: officials http://www.cnn.com/2003/HEALTH/04/24/sars.death/ Date: April 25, 2003 Date accessed: March 21, 2020

